# Chikungunya’s return amidst 2024 dengue season: Implications for future outbreaks in Bangladesh

**DOI:** 10.1101/2024.12.23.24319578

**Authors:** Anamul Hasan, Md. Fahad Zamil, Afrida Tabassum Trina, Mohammad Sharif Hossain, Sajia Afreen, Dilruba Ahmed, Mohammad Shafiul Alam

**Affiliations:** Infectious Diseases Division, International Centre for Diarrheal Disease Research, Bangladesh (icddr,b), Dhaka, Bangladesh; Clinical and Diagnostic Services, International Centre for Diarrheal Disease Research, Bangladesh (icddr,b), Dhaka, Bangladesh

**Keywords:** Chikungunya, Bangladesh, Dhaka, Re-emergence, Outbreak

## Abstract

Chikungunya, an arboviral disease transmitted by *Aedes* mosquitoes, shares clinical similarities with dengue, but is distinguished by prolonged joint pain. Following a major outbreak in Bangladesh in 2017, chikungunya nearly vanished from its territory. The study participants were recruited prospectively following specific inclusion criteria and obtained written informed consent. Out of 1280 febrile individuals screened, 569 met the criteria of fever onset within 2-5 days, accompanied by symptoms such as headache, myalgia, bone and joint pain, rash, nausea, vomiting, or diarrhea. Of these, 474 underwent real-time RT-PCR testing. Among the samples tested, 213 were PCR-positive for at least one arbovirus. Chikungunya cases totaled 55, including 7 coinfections (6 with DENV, and the first documented CHIKV-ZIKV coinfection in Bangladesh). No infections were reported from January to August, with a peak in October and November. Most CHIKV infections (72.7%) had moderate to high viral loads, with common symptoms of joint pain, myalgia, and headaches. The resurgence of Chikungunya in late 2024 underscores the potential for a major outbreak in 2025, necessitating proactive measures to mitigate public health impact and ensure a robust response to this re-emerging threat.

## Introduction

Chikungunya, transmitted by *Aedes* mosquitoes, shares common vectors with dengue and presents with similar clinical features, including fever, joint and muscle pain, and rash. However, chikungunya is distinguished by its prolonged joint pain, which may persist significantly longer than in cases of dengue (Furuya-Kanamori et al., 2016). Co-infection with chikungunya virus (CHIKV) alongside other arboviruses, such as dengue (DENV) or Zika virus (ZIKV), poses a significant risk to public health, creating a prolonged burden on both patients and healthcare resources (Taraphdar et al., 2022, Carrillo-Hernández et al., 2018).

Chikungunya outbreaks was first noticed in 2008 in the northwest part of Bangladesh, affecting 39 people (Icddr, 2009), followed by 196 cases in Nababgonj and Dohar sub-districts of Dhaka in 2011, with sporadic cases reported in 2013, 2014, and 2015, leading to a significant outbreak in December 2016 (Alam, 2018). The most severe outbreak of chikungunya in Bangladesh occurred between April and September 2017, outranking previous incidents, with over 13,000 clinically confirmed patients reported in Dhaka alone and overall cases documented across 23 districts nationwide (Anwar et al., 2020).

In 2023, Bangladesh faced the largest and deadliest dengue outbreak since the virus reemerged in the country two decades ago, with 1,705 fatalities marking the highest case fatality rate (0.5%) due to dengue globally for that year (Hasan et al., 2024b). Concurrently, five cases of ZIKV transmission were diagnosed as well (Hasan et al., 2024a). Given this context and the rising concern over arboviral infections, there is a critical need to detect and monitor these viruses accurately. Our study aimed to simultaneously detect the presence of DENV, CHIKV, and ZIKV using a real-time multiplex RT-PCR system, considering the relevant symptomatology, to effectively address these pressing public health challenges.

## Materials and Methods

The study participants were enrolled after meeting the inclusion and exclusion criteria and providing informed written consent or assent. The inclusion criteria included onset of fever within 2 – 5 days, along with one of the following clinical features, e.g. headache, myalgia, bone and joint pain, rash, nausea and vomiting, or diarrhea were recruited prospectively for a diagnostic evaluation study. The study was conducted in the Dhaka city corporation areas, Bangladesh. Individuals reporting to the icddr,b diagnostic facilities of for dengue tests were considered for primary screening. Suspected cases aged between 5 and 65 years were screened. The exclusion criteria included individuals outside this age range, those with severe illness, or those with conditions that contraindicated invasive sample collection. After meeting the inclusion and exclusion criteria, if the participant provided informed written consent or assent was enrolled.

The study was carried out from January to November 2024. Using aseptic techniques and universal precautions, approximately 3ml of venous blood was drawn from each febrile patient into serum-separator tubes (SST). Serum samples were isolated by centrifugation and aliquoted into 1.5ml Eppendorf tubes. Each sample initially underwent Dengue NS1 rapid diagnostic testing (RDT) with the Bioline Dengue NS1 antigen kit (Abbott, Ingbert, Germany, Cat# 11FK50).

Viral RNA was extracted from 140μL of serum using the QIAmp Viral RNA Mini kit (Qiagen, Hilden, Germany, Cat# 52906) following the manufacturer’s protocol. RNA samples were then cryo-preserved at -80□C until batchwise real-time RT-PCR was conducted. Arboviral RNA (DENV, CHIKV, or ZIKV) was detected using the VIASURE Zika, Dengue & Chikungunya Real Time RT-PCR Detection Kit (CerTest Biotec, Zaragoza, Spain, Cat# VS-ZDC106L) as per the manufacturer’s instructions. Thermal cycling was performed on the CFX-96 real-time PCR detection system (Bio-Rad, Hercules, CA, USA).

Raw data were entered into Google spreadsheets, and analyzed and curated using STATA (v.15) (StataCorp LLC, College Station, TX, USA). Separate datasets were maintained for different case types, with thorough scrutiny to ensure data integrity, eliminate duplicates, and correct inaccuracies and inconsistencies. These datasets were eventually merged into a final comprehensive dataset, with individual datasets retained as backups. Graphical presentations were created using GraphPad Prism (v.9) (GraphPad Software Inc., La Jolla, CA, USA).

## Result

Initially, a comprehensive screening procedure was conducted on a total of 1,280 individuals. Out of these, 569 participants met the pre-defined inclusion criteria and provided written informed consent or assent, thereby officially enrolling in the study. Regardless of the results from the Dengue NS1 RDT, the RT-PCR analyses were subsequently performed on 474 samples selected using a convenient sampling technique, resulting in 213 as tested PCR-positive for at least one of the three targeted arboviruses— DENV, CHIKV, or ZIKV. The remaining 261 samples were confirmed as negative for these viruses. Out of 55 CHIKV cases, only 7 cases had a history of co-infection with either DENV (n = 6) or ZIKV in single case (**Figure 1.A**).

**Figure 1.**
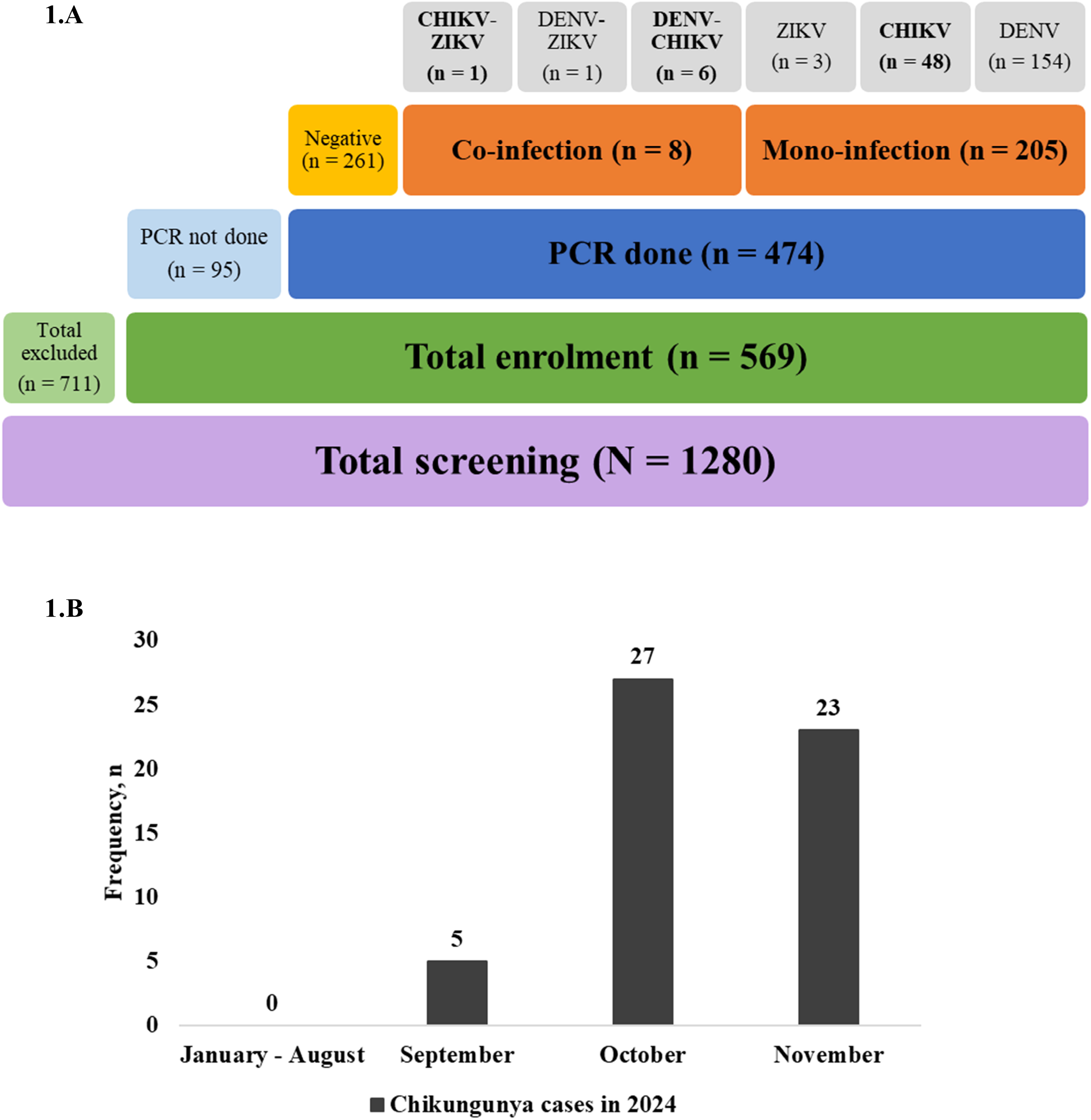
Screening and enrollment data for viral infections. 1.A) Flowchart showing the screening and enrollment process, with totals for screened, excluded, enrolled, PCR tested, and infection outcomes. 1.B) Bar graph depicting monthly Chikungunya case frequencies in 2024 (December not included), highlighting a peak in October and November.

Analysis of the monthly distribution reveals no reported chikungunya cases from January to August in 2024. However, a notable increase was observed from September onwards, reaching a peak in October, and slightly decreasing in November. This trend indicates a significant rise in Chikungunya incidence during the late monsoon period (**Figure 1.B**).

The cohort of 55 individuals diagnosed with CHIKV infection consisted of a disproportionate ratio of male and female (1.6:1), while a significant number of participants were predominantly aged between 18-30 years (n = 19). Most participants experienced fever onset within 2 days (n = 34), indicating a rapid progression of symptoms following infection. The Ct values of RT-PCR, inversely indicating viral load, showed that 72.7% of cases (n = 40) had moderate to higher viral loads (Ct < 30). (**Table 1**).

**Table 1.** Clinico-demographic criteria of CHIKV infected participants (n = 55)

| Criteria | Sub-group | Frequency (n) |
| --- | --- | --- |
| Gender | Male | 34 |
|  | Female | 21 |
| Age group | 5 – 17 | 6 |
|  | 18 – 30 | 19 |
|  | 31 – 40 | 8 |
|  | 41 – 50 | 5 |
|  | 51 – 60 | 13 |
|  | Above 60 | 4 |
| Onset of fever (days) | 2 | 34 |
|  | 3 | 18 |
|  | 4 | 3 |
| Symptoms | Bone and joint pain | 51 |
|  | Myalgia | 48 |
|  | Headache | 45 |
|  | Nausea and vomiting | 35 |
|  | Fatigue | 32 |
|  | Retro-orbital pain | 27 |
|  | Rash | 15 |
|  | Itching | 9 |
|  | Eye inflammation | 9 |
|  | Diarrhea | 9 |
|  | Loss of appetite | 8 |
| Ct value of RT-PCR<br>(45 cycles) | Below 20 | 10 |
|  | 20 – 25 | 14 |
|  | 25 – 30 | 16 |
|  | 30 - 40 | 15 |

The most commonly reported symptoms include bone and joint pain, which was observed in a majority of the cases (n = 51), followed by myalgia (n = 48), headaches (n = 45), and nausea and vomiting (n = 35). Notably, one participant experienced gum bleeding, while another suffered from lymph node swelling (not shown in table).

## Discussion

The CHIKV infection is often underdiagnosed and underreported due to its symptomatic similarities to DENV, leading to limited molecular diagnostic testing for CHIKV in Bangladesh. A national seroprevalence survey conducted shortly before the 2017 epidemic found a 2.4% seropositivity rate for CHIKV, and a spatial prediction model estimated that nearly 5 million people in Bangladesh had previously been infected. Since the large-scale epidemic in 2017, no major chikungunya outbreaks have been reported so far (Allen et al., 2024). However, the recent resurgence of the DENV-2 serotype in 2023 (Hasan et al., 2024b), with its continued predominance in ensuing year of 2024, raises concerns about a potential increase in chikungunya cases. Without prompt and early intervention, chikungunya could pose a substantial public health concern in the upcoming year of 2025.

We reported the first case of CHIKV-ZIKV co-infection from Bangladesh, to our knowledge. The clinical implications of CHIKV-ZIKV co-infection are somewhat rare and complex, leading to significant challenges in diagnosis and treatment. Such co-infections often result in more severe disease manifestations, including prolonged fever, increased joint pain, and a heightened risk of neurological complications. Despite its low global prevalence rate of 1.0%, the potential severity underscores the need for vigilant monitoring and advanced clinical management (Ahmed et al., 2024).

The incidence of arboviral infections is significantly influenced by various seasonal factors like temperature, humidity, and rainfall, as well as human behavior patterns, their migration for urbanization (Hasan et al., 2024b). The possible correlation between the re-emergence of CHIKV infections and the late monsoon pattern in 2024 might be orchestrated in the massive CHIKV outbreak in the ensuing year of 2025 in Bangladesh (Subhadra et al., 2021). A similar event was observed during the dengue upsurge in Bangladesh in 2023, which occurred following the late monsoon period in 2022 (Hasan et al., 2024b).

There are some limitations to consider. This study was not a population representative survey and did not analyze the risk of hospitalization and any type of hemorrhagic fever in case of secondary infection.

The identification of chikungunya cases was immediately notified to the Directorate General of Health Services of Bangladesh as per the Communicable Diseases (Prevention, Control and Eradication) Act, 2018 (DGHS, 2018). The information was also shared with the corresponding city corporations for their actions.

## Data Availability

All data produced in the present study are available upon reasonable request to the authors.

## Acknowledgements

We acknowledge with gratitude the commitment of CTK Biotech Inc., USA to its research efforts. We are also grateful to the Governments of Bangladesh and Canada for providing core/unrestricted support. Our special gratitude goes to the field study team for their relentless support to make it possible.

## Ethical approval

The study was approved by the institutional ethical review committee (ERC) of the International Center for Diarrheal Disease Research, Bangladesh (icddr,b). The institutional review board (IRB) approved the study with protocol no PR-23080. The corresponding ethical consent/assent has been obtained from the participants.

## Author contributions

Conception: MSA.

Investigations: ATT, MFZ, SA and AH.

Methodology: AH, DA and MSA.

Data Analysis: ATT, MFZ, MSH, SA and AH.

Validation: MSH and MSA.

Supervision: DA and MSA.

Writing Original: AH.

Writing-review & editing: MSA.

All authors reviewed and approved the final version of the correspondence.

